# Validation of a NGS panel, with automated analysis, designed for detection of medically actionable tumor biomarkers for Latin America

**DOI:** 10.1101/2021.03.19.21253988

**Authors:** Mauricio Salvo, Evelin González-Feliú, Jessica Toro, Iván Gallegos, Ignacio Maureira, Nicolás Miranda González, Olga Barajas, Eva Bustamante, Mónica Ahumada, Alicia Colombo, Ricardo Armisén, Camilo Villamán, Carolina Ibañez, María Loreto Bravo, Verónica Sanhueza, Loreto Spencer, Gonzalo de Toro, Erik Morales, Carolina Bizama, Patricia García, Ana María Carrasco, Lorena Gutiérrez, Justo Lorenzo-Bermejo, Ricardo A. Verdugo, Katherine Marcelain

**Affiliations:** Department of Basic and Clinical Oncology, Faculty of Medicine, Universidad de Chile, Santiago, Chile; Department of Pathology, Hospital Clínico de la Universidad de Chile, Santiago, Chile; Department of Medical Technology, Faculty of Medicine, Universidad de Chile, Santiago, Chile; Department of Internal Medicine, Hospital Clínico Universidad de Chile. Santiago, Chile; Fundación Arturo López Pérez, Santiago, Chile; Centro de Genética y Genómica, Instituto de Ciencias e Innovación en Medicina, Facultad de Medicina, Clínica Alemana Universidad del Desarrollo, Santiago, Chile; Hematology & Oncology Department, Faculty of Medicine, Pontificia Universidad Católica de Chile (PUC). Santiago, Chile; Servicio de Anatomía Patológica, Hospital Padre Hurtado, Santiago, Chile; Servicio de Anatomía Patológica, Hospital Clínico Regional Guillermo Grant Benavente, Concepción, Chile; School of Medical Technology, Universidad Austral de Chile at Puerto Montt, Puerto Montt, Chile; Servicio de Anatomía Patológica, Hospital Regional de Talca, Talca, Chile; Departament of Preclinical Sciences, Faculty of Medicine, Universidad Católica del Maule, Talca, Chile; Department of Pathology, Faculty of Medicine, Pontificia Universidad Católica de Chile, Santiago, Chile; Department of Pathology, Hospital San Juan de Dios. Santiago, Chile; Institute of Medical Biometry and Informatics, Universität Heidelberg, Germany; Human Genetics Program, ICBM, Faculty of Medicine, Universidad de Chile. Santiago, Chile

**Keywords:** NGS-panel, oncology, target therapies, predictive biomarkers, automated bioinformatic analysis, somatic variants, Latin America

## Abstract

The genomic characterization of solid tumors and a rapidly growing repertoire of target drugs are revolutionizing cancer treatment. Next-generation sequencing (NGS) panels are progressively used in clinical practice for target therapy in high-income countries. In contrast, limited access to tumor sequencing, among other barriers, precludes precision cancer treatment in low- and middle-income countries. To build towards the implementation of precision oncology in Chile and Latin America, we designed a 25-gene panel that contains predictive biomarkers for currently or near-future available therapies in Latin America. Library preparation was optimized to account for DNA integrity variability in Formalin-Fixed Paraffin-Embedded (FFPE) tissue. The bioinformatic pipeline removes FFPE-induced artifacts and known germline variants; while identifying possible discrepancies in somatic mutations due to Latin Americans’ underrepresentation in the reference genome databases. Analytic sensitivity and accuracy were assessed using commercial standard controls for FFPE DNA and for germline BRCA1 and BRCA2 mutations, which are biomarkers for PARP inhibitors. Our panel detects small insertions and deletions and single nucleotide variants (SNVs) with 100% sensitivity and specificity down to allelic frequencies of 0.05, and with 100% between-run and within-run reproducibility for non-synonymous variants. The workflow was validated in 265 clinical samples, including breast, colorectal, gastric, ovarian, and gallbladder tumors and blood, leading to identifying 131 actionable variants. Therefore, this NGS panel constitutes an accurate and sensitive method for routine tumor biopsies that could replace multiple non-NGS assays and costly large NGS panels in the Latin American clinical context. The proposed streamlined assay and automated analysis are expected to facilitate the implementation of precision medicine in Latin America.

## Introduction

Over the last few decades, molecular pathology has substantially advanced thanks to exponential genetic sequencing technology growth. The introduction of next-generation sequencing (NGS) opened the doors to high-throughput, multi-gene, massive data collection. This tool’s ability to sequence more, faster, and at a reduced cost has made it attractive for many clinical research applications. In cancer, using this technology to interrogate solid tumor samples has propelled a massive characterization of genes involved in the disease^1,2^. This rise in “oncogenomics” has been accompanied by an increase in cancer drug approval and development^3,4^. Identifying tumor-specific genetic signatures and their correlation to treatment outcome have evolved into a strategy coined ‘precision medicine’, a new diagnose and treat process in cancer based on approved genomic biomarkers^4^.

In Latin America and the Caribbean, 1.4 million new cancer cases were estimated to occur in 2018, while mortality rates vary among and within the region^5,6^. In Latin America, the most common types of cancer with the highest incidence are prostate (Age Standardized Rate (ASR) 60.4), breast (ASR 56.8), colorectal (ASR 18.6), cervix uteri (ASR 15.2), lung (ASR 13.1), and stomach (ASR 9.5) cancers^1^. Overall, estimated age-standardized cancer incidence rates in Latin America are lower than those reported in North-America and some European countries; however, the region exhibits higher mortality rates^7^. This paradox reflects the disparities in early diagnosis and treatment opportunities in the region.

While in high and medium-income countries, precision medicine is making its way into standard cancer treatment, improving survival and investigational drug trial success for many patients, a combination of factors prevent this helpful tool from becoming accessible to most of the world population. In many countries, approved and available assays are often designed to meet foreign standards and may even require international sample shipping and/or data processing. These standard designs often interrogate parts of the genome that may be clinically irrelevant for some locations where not all treatment options are available, significantly inflating sequencing costs. Besides, the absence of automated clinician-ready reporting for many of these approved panels creates another major cost and obstacle to widespread implementation. As a result, NGS-based oncology panels do not appear cost-effective solutions to many governments and are not getting implemented in health and insurance systems despite local sequencing capabilities. This scenario creates an urgent need for customized validated solutions and data interpretation in a clinical environment^8^.

In addition, an important caveat to interpreting Latin American cancer patient’s genetic data is the under-representation of Latin American individuals in the global resources characterizing the frequency of both germinal and cancer genome variants. Great cancer genomics efforts, like TCGA and ICGC, are deprived of minorities (including subjects of Hispanic ethnicity^35^), limiting the capacity to describe somatic mutations with a prevalence below 10% and overcome the somatic background mutation frequency in specific ethnic groups^36^. For instance, the average Amerindian ancestry in cancer patients across all cohorts in TCGA is about 4%^34,35^. Also, the Latin American population is under-represented in germline variant repositories, which may induce a false categorization and overestimation of somatic variants^9–11^. Thus, an additional blood sample should accompany the tumor sample, increasing the sequencing costs.

To address these challenges, our team designed, optimized, and validated a hybridization-based target enrichment workflow with multiple automated analyses capable of detecting variants in 25 genes associated with approved and standard of care target therapies across multiple sample types. Although this panel was designed to meet current and near-future Chilean precision oncology needs, we expect the panel and workflow to be relevant to other countries in the region. We validated this workflow locally using breast, colorectal, gastric, ovarian, pancreatic, and gallbladder tumor tissue samples and report the ability to detect single nucleotide variants (SNVs) and small insertions and deletions with 100% sensitivity and specificity. Additionally, 100% reproducibility was obtained for non-synonymous variants between and within runs. Finally, to address the shortage of health professionals trained in bioinformatics, the entire workflow, including quality control of sequencing data and calling for somatic variants, was automated and made available.

## Materials and Methods

### Panel Design

We designed and constructed an NGS panel for predictive biomarkers in solid tumors that target hotspots, selected exons, or complete coding regions for 25 genes. We refer to this panel, plus its associated workflow and analysis, as TumorSec™. For selecting targeted regions, biomarker genes classified with evidence 1, 2, 3a, 3b, R1, and R2 were selected for solid tumors in the OncoKB database (www.oncokb.org)^33^. Next, biomarker mutations with level of clinical evidence A, B, and C were selected in the Clinical Interpretation of Variants in Cancer, CiVic database (https://civicdb.org/home)^32^ and manually curated. Biomarkers were selected based on their level of evidence and incidence of the targeted tumor in Latin America. *TP53* and *ARID1A* complete coding regions were incorporated, as they contain prognosis and predictive chemotherapy biomarkers. The complete list of genes and drug associations is provided in Table 1.

**Table 1.** Genes included in the panel and their therapy association.

| GENE | Target Region | Drugs | Tumor Type | Highest level of evidence* |
| --- | --- | --- | --- | --- |
| AKT1 | all exons | AZD-5363 | Breast Cancer<br>Ovarian Cancer<br>Endometrial Cancer | B |
| ALK | Hot spots | Ceritinib<br>Crizotinib<br>Alectinib<br>Brigatinib<br>Lorlatinib | Non-Small Cell Lung Cancer | A |
| ARID1A | all exons | ATM inhibitors<br>Erlotinib<br>Chemotherapy | Breast Cancer<br>Ovarian Cancer | C |
| BRAF | all exons | Encorafenib + Cetuximab<br>Vemurafenib<br>Dabrafenib<br>Trametinib + Dabrafenib<br>Cobimetinib + Vemurafenib<br>Trametinib<br>Encorafenib + Binimetinib<br>Vemurafenib + Cobimetinib,<br>Trametinib + Dabrafenib<br>Vemurafenib + Cobimetinib<br>Encorafenib + Panitumumab | Colorectal Cancer<br>Melanoma<br>Non-Small Cell Lung Cancer<br>Anaplastic Thyroid Cancer<br>Hairy Cell Leukemia<br>Pilocytic Astrocytoma<br>Ganglioglioma<br>Pleomorphic Xanthoastrocytoma | A |
| BRCA1 | all exons | Olaparib | Ovarian Cancer<br>Peritoneal Serous Carcinoma<br>Breast Cancer<br>Prostate<br>Ovary/Fallopian Tube | A |
| BRCA2 | all exons | Olaparib | Ovarian Cancer<br>Peritoneal Serous Carcinoma<br>Breast Cancer<br>Prostate<br>Ovary/Fallopian Tube | A |
| Cdk4 | all exons | Palbociclib<br>Abemaciclib | Dedifferentiated Liposarcoma<br>Well-Differentiated Liposarcoma | A |
| EGFR | Hot spots | Erlotinib<br>Afatinib<br>Osimertinib<br>Gefitinib<br>Dacomitinib | Non-Small Cell Lung Cancer | A |
| ERBB2 | Hot spots | Trastuzumab | Breast Cancer | A |
| ESR1 | Hot spots | Palbociclib | Breast Cancer | B |
| IDH2 | Hot spots | Enasidenib | Acute Myeloid Leukemia | A |
| KIT | Hot spots | Sunitinib<br>Imatinib<br>Regorafenib<br>Sorafenib | Gastrointestinal Stromal Tumor | A |
| KRAS | all exons | Cetuximab<br>Erlotinib<br>Regorafenib<br>Selumetinib<br>Irinotecan | Colorectal Cancer<br>Pancreatic Cancer<br>Non-Small Cell Lung Cancer | A |
| MET | Hot spots | Foretinib, Rilotumumab | Papillary Renal Cell Carcinoma<br>Melanoma<br>Non-Small Cell Lung Cancer | B |
| MTOR | all exons | mTOR inhibitors | Renal Cell Carcinoma | B |
| NRAS | all exons | EGFR inhibitors | Colorectal Cancer | A |
| PDGFRA | Hot spots | Imatinib<br>Sunitinib | Gastrointestinal Stromal Tumor | A,B |
| PI3KCA | all exons | Buparlisib<br>Serabelisib<br>Alpelisib<br>Copanlisib | Breast Cancer | A |
| PTCH1 | all exons | Vismodegib | Medulloblastoma<br>Skin cancer | B |
| PTEN | all exons | Everolimus | Various | B |
| ROS1 | Hot spots | Crizotinib | Non-Small Cell Lung Cancer | C |
| SMO | Hot spots | Vismodegib | Basal Cell Carcinoma | B |
| TP53 | all exons | Prognosis | Various | A |
| TSC1 | all exons | MTOR inhibitors, Everolimus | Renal Cell Carcinoma, Bladder Cancer | A |
| TSC2 | all exons | MTOR inhibitors | Central Nervous System<br>Renal Cell Carcinoma | A, B |

\*According to AMP/ASCO/CAP. Level **A**: FDA-approved therapy included in professional guidelines; **B**: Well-powered studies with consensus from experts in the field; **C**: FDA-approved therapies for different tumor types or investigational therapies. Multiple small published studies with some consensus (Li, MM et al. J Mol Diagn 2017). The highest level of evidence is shown. Some biomarkers may have additional indications with lower levels of evidence for different cancer types or protocols.

Synthesis of the soluble, biotinylated probe library was done on the NimbleGen cleavable array platform (SeqCap EZ Choice; Roche/NimbleGen, Basel, Switzerland). The probe design was optimized using the NimbleDesign software utility, which adjusts individual probe lengths based on melt temperatures and homopolymer repeat length (NimbleGen, Roche; http://sequencing.roche.com/products/software/nimbledesign-software.html).

### Sample Information

The study was approved by the “Comité de Ética Científico del Servicio Metropolitano de Salud Oriente”, “Comité Ético Científico o de Investigación del Hospital Clínico de la Universidad de Chile” (approval number N°17-18), and the “Comité de Ética de Investigación en Seres Humanos de la Facultad de Medicina de la Universidad de Chile”. Colorectal and gastric cancer samples were obtained from the “Biobanco de Tejidos y Fluidos de la Universidad de Chile”. To capture real world heterogeneity in sample quality, breast, ovary and gallbladder FFPE tissue samples were collected from the pathology services from several sites along the country (Fundación Arturo López Pérez, Clínica Dávila, Clínica Indisa, Red UC Christus, Biobanco de Tejidos y Fluidos, Hospital Padre Hurtado, Hospital Regional de Concepción, Hospital Regional de Talca, Hospital de Puerto Montt, Hospital San Juan de Dios, Hospital Santiago Oriente Doctor Luis Tisné Brousse, Instituto Nacional del Cáncer, Hospital del Salvador, Hospital Regional de Coquimbo, Hospital Regional de Arica, Hospital Clínico San Borja Arriarán). All individuals were informed about the planned molecular testing, and signed an informed consent in advance for the retrieval and processing of their data and samples.

A total of 176 tumor tissue samples were sequenced for this study. Nineteen were freshly frozen (FF): 13 colorectal and six breast; 157 were formalin-fixed paraffin-embedded (FFPE) blocks: 9 breast, 71 ovary, one gastric, 36 gallbladder, and 40 colorectal tumors. Additionally, DNA from 89 whole blood or buffy coat samples were sequenced: 7 from colorectal and 72 from breast cancer patients.

### DNA Extraction, quantification, and Sample Quality Control

DNA from Frozen Tissues was extracted using the QIAamp DNA Mini Kit (Qiagen). FFPE tissue DNA was extracted using GeneJet FFPE DNA Purification Kit and RecoverAll(™) Total Nucleic Acid Isolation (Invitrogen, Thermo Fisher Scientific), following the manufacturer’s instructions, with overnight lysis instead of the suggested 1-2 hour for FFPE tissue. Germline DNA was purified from whole blood samples using the Wizard^®^ Genomic DNA Purification Kit (Promega), according to manufacturer’s protocol.

Purified DNA was quantified using the Qubit(™) dsDNA HS Assay and Quant-IT(™) Picogreen^®^ dsDNA Reagent Kit (Invitrogen, Thermo Fisher Scientific). The purity of DNA was assessed by measuring the 260/280 nm absorbance ratio. For FFPE samples, fragment length and degradation were assessed using the HS Genomic DNA Analysis Kit (DNF-488) in a Fragment Analyzer (Agilent, formerly Advanced Analytical). DNA ranged from >1000bp to less than 200bp (Supplementary Figure 1).

### Library preparation

100-150ng of DNA (Blood and Frozen Tissues) and 200ng of DNA (FFPE) were used as input for sequencing library preparation. NGS libraries were prepared using KAPA HyperPlus Library Preparation Kit (Kapa Biosystems), DNA was incubated with the HyperPlus “Frag Enzyme mix” (KAPA Biosystems) with modifications for FFPE DNA samples. End repair and phosphorylation were performed according to the manufacturer’s protocols. Ligation of barcoded adaptors reaction was modified for FFPE DNA. We performed a double size selection in libraries prepared with DNA from frozen tissue and blood and a single size selection in libraries prepared with DNA from FFPE. Libraries were quantified using the QubitTM dsDNA HS Assay Kit (Invitrogen) and Quant-IT^™^ Picogreen^®^ dsDNA Reagent Kit (Invitrogen). The quality of the amplified library was checked by measuring the 260/280 absorbance ratio and fragment’s length, using the HS NGS Analysis Kit (DNF-474) in a Fragment Analyzer (Agilent, formerly Advanced Analytical).

### Target Enrichment

Prepared DNA libraries (1200ng total mass) were captured by hybridization probes (Roche NimbleGen SeqCap EZ). The number of samples used for pre-capture multiplexing was based on sample type: six were pooled for FFPE and blood samples, while fresh frozen tumor samples were pooled in reactions of four. As with the sample libraries, captured libraries were assessed for concentration and size distribution to determine molarity.

### Sequencing run Set-up

Libraries were diluted to a concentration of 4 nmol/L and process for sequencing, according to the manufacturer’s instructions (Illumina, San Diego, CA). The final captured library concentration for sequencing was 9.4 pM - 9.5 pM. Libraries were sequenced in an Illumina^®^ MiSeq System using paired-end, 300 cycles (MiSeq Reagent Kits v2, Illumina^®^).

### Control Samples

Three reference standard DNA samples from Horizon Discovery (Cambridge, United Kingdom) were used as positive controls for variant calling: HD200 (FFPE somatic), HD793, and HD794 (germline BRCA1/2 variants).

### Bioinformatic Analyses

TumorSec’s bioinformatic package is a compilation of open-source programs that are executed sequentially and automatically from a locally developed bash program to call out somatic variants (SNVs and indels) from Illumina sequencing output from FFPE or fresh tumor samples, without the need for germline sample to detect somatic variants (Figure 1).

**Figure 1.**
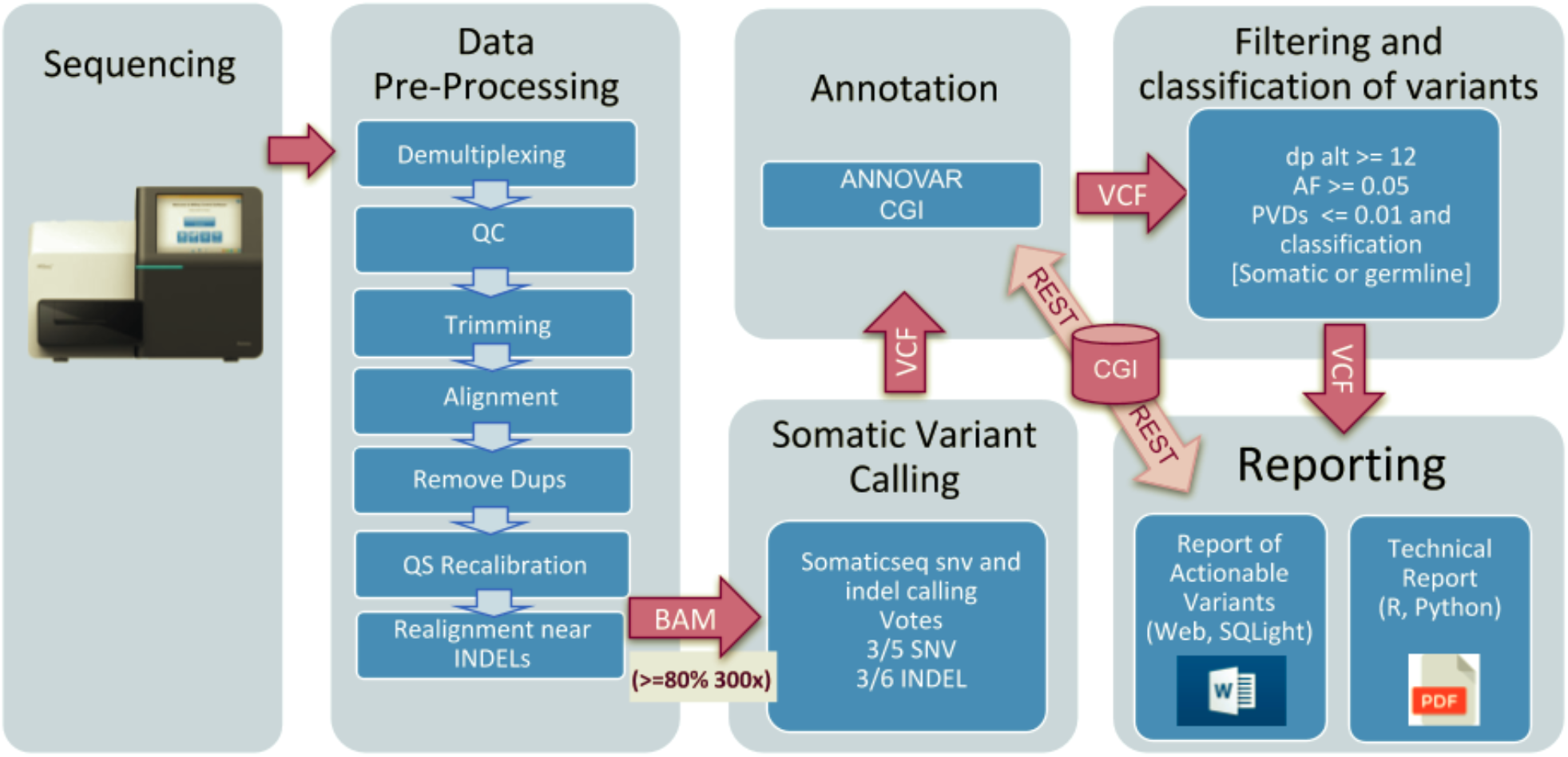
General workflow of the bioinformatic pipeline for identifying somatic variants. Gray boxes correspond to automated processes. Blue squares show the main sub-processes. Key input and output files are shown in arrows.

#### Data pre-processing

The bioinformatic workflow starts with the demultiplexing of the basecall files generated by the sequencing cycle using the Illumina bcl2fastq v2.20.0 program, generating sample-specific FASTQ files. Filtering of reads and base correction is done with the fastp v0.19.11 tool, which implements a sliding window algorithm to remove low-quality bases at the ends of each read, removing reads with less than 50 nucleotides or with a Phred quality score under 20. The filtered reads align with the reference genome GRCh37/hg19 using Burrows-Wheeler Alignment mem (BWA mem v0.7.12) with the default parameters. Subsequently, the MarkDuplicates tool of Picard v2.20.2-8 is applied to identify pairs of reads that have originated from the same DNA fragment product of the clustering or PCR artifact. Then, to reduce the number of mismatches to the reference genome, the reads are realigned with RealignerTargetCreator and IndelRealigner from GATK v3.8 ^12^. Finally, the quality scores are re-calibrated with the combination of GATK’s BaseRecalibrator and PrintReads tools ^12^.

#### Somatic Variant Calling and annotation

The SomaticSeq v.3.3.0 program was used to call the variants in single-mode using only the tumor sequence data^13^. This tool maximizes the sensitivity by combining the result of five next-generation variant SNV callers - Mutect2 ^14^, VarScan2 ^15^, VarDict ^16^, LoFreq ^17^, and Strelka ^18^ - adding Scalpel for indels. A minimum allelic frequency of 0.1 was set for variants search, and the rest of the parameters are integrated by default. Each variant caller generates a separate VCF file, and SomaticSeq combines these files to detect consensus variants. The reported SNVs are identified by at least 3 out of 5 SNV callers, and the reported indels by at least 3 out of the six callers. The consensus variants obtained by SomatiSeq were annotated using the Cancer Genome Interpreter (https://www.cancergenomeinterpreter.org/), reporting validated oncogenic alterations, driver mutations, and also associates genomic biomarkers of drug response ^19^. Additionally, the variants were annotated with ANNOVAR^20^ using RefGene, GnomAD v2.1.1 (genome and exome), ESP6500, ExAC v0.3, 1000 Genomes phase 3, CADD v1.3, dbSNP v150, COSMIC v92, and CLINVAR.

#### Variant filtering and sequence quality reporting

Variants with allele frequencies greater than 0.5 and with an altered allele depth ≥12 reads were selected. These thresholds were established as our Limit of Detection (LOD) for the NGS TumorSec panel following the recommendations of the Association for Molecular Pathology and College of American Pathologists^21^. Polymorphisms were eliminated, discarding alleles with a frequency greater than 0.01, reported in 1000 Genomes, ESP6500, GnomAD or ExAC^20^. Filtering was extended to include all under-represented populations that have information in GnomAD and ExAC. Additionally, a local repository was built using germline data from this and other related works. Finally, since no noncoding or synonymous mutations have been reported as biomarkers of therapeutic response in cancer, only mutations that produce a protein change (non-synonymous) were selected for reporting.

The bioinformatics pipeline is executed automatically, creating pdf reports that allow an easy view of quality metrics per sample. For this purpose, the programs FastQC v 0.11.8, Qualimap v2.2.2a, Mosdepth v0.2.5, and MultiQC v1.8 are executed between the pre-processing of the bioinformatics workflow, and the quality metrics are obtained in a pdf file. The main metrics are the number of initial raw reads, the percentage of filtered reads, the duplication rate, the number of reads on target regions, the average depth on-target regions, the uniformity percentage, and the ratio of on-target regions with a minimum coverage of 100X to 500X. For variant calling and annotation, we set a coverage threshold for FFPE and FF of 300X in at least 80% of target regions. The bioinformatic pipeline, user manual, and tutorials can be found in the GitHub repository called Pipeline-TumorSec (https://github.com/u-genoma/Pipeline-TumorSec).

#### Germline variant calling

Data were pre-processed following the same protocol for somatic variants observed in Figure 1. From the BAM files per sample, variant calls were made using the GATK HaplotypeCaller tool, which realigns each haplotype to the reference (hg19) using the Smith-Waterman algorithm to identify SNPs and indels. A minimum confidence threshold of 30 was set for variant calling. Additionally, we set a coverage threshold of 200X in at least 80% of target regions. Finally, a variant calling hard-filter for SNPs and indels was applied separately following GATK recommendations^22^.

## Results

### Panel Design and Sequencing metrics

A total of 25 genes were included in this target enrichment panel, covering 98 kb of sequence length. Design details broken down by gene are shown in Table 1. The decision to include all exons or only hot spots per gene was based on the number of relevant genomic features. 79% (15/19) of fresh frozen, 69% (108/157) of FFPE, and 89% (79/89) of blood samples processed passed the sequencing quality threshold capturing a minimum of 80% of target regions at 300X of depth. A summary of relevant sequencing metrics for all 202 passed samples is shown in Table 2. FFPE samples showed a high percentage of duplicate and off-target reads. Uniformity was > 90% for all sample types and >90% of targeted regions had ≥300x coverage (Figure 2A). Sequencing coverage across the targeted regions was >90% (Figure 2B).

**Table 2.** Sequencing metrics (median) for all samples that passed QC.

|  | Sample Type |  |  |
| --- | --- | --- | --- |
| Sequencing Metric | FFPE<br>(n=108) | Blood<br>(n=79) | Fresh Frozen<br>(n=15) |
| % Duplicated reads | 34.4 | 9.5 | 9.15 |
| % of reads in target region | 58.8 | 69.1 | 65.8 |
| Average coverage in target region | 1,133.8 | 798.5 | 789.1 |
| % Uniformity | 93.7 | 96.0 | 91.1 |
| % of target regions with minimum coverage of 300X | 99.0 | 97.4 | 90.0 |

**Figure 2.**
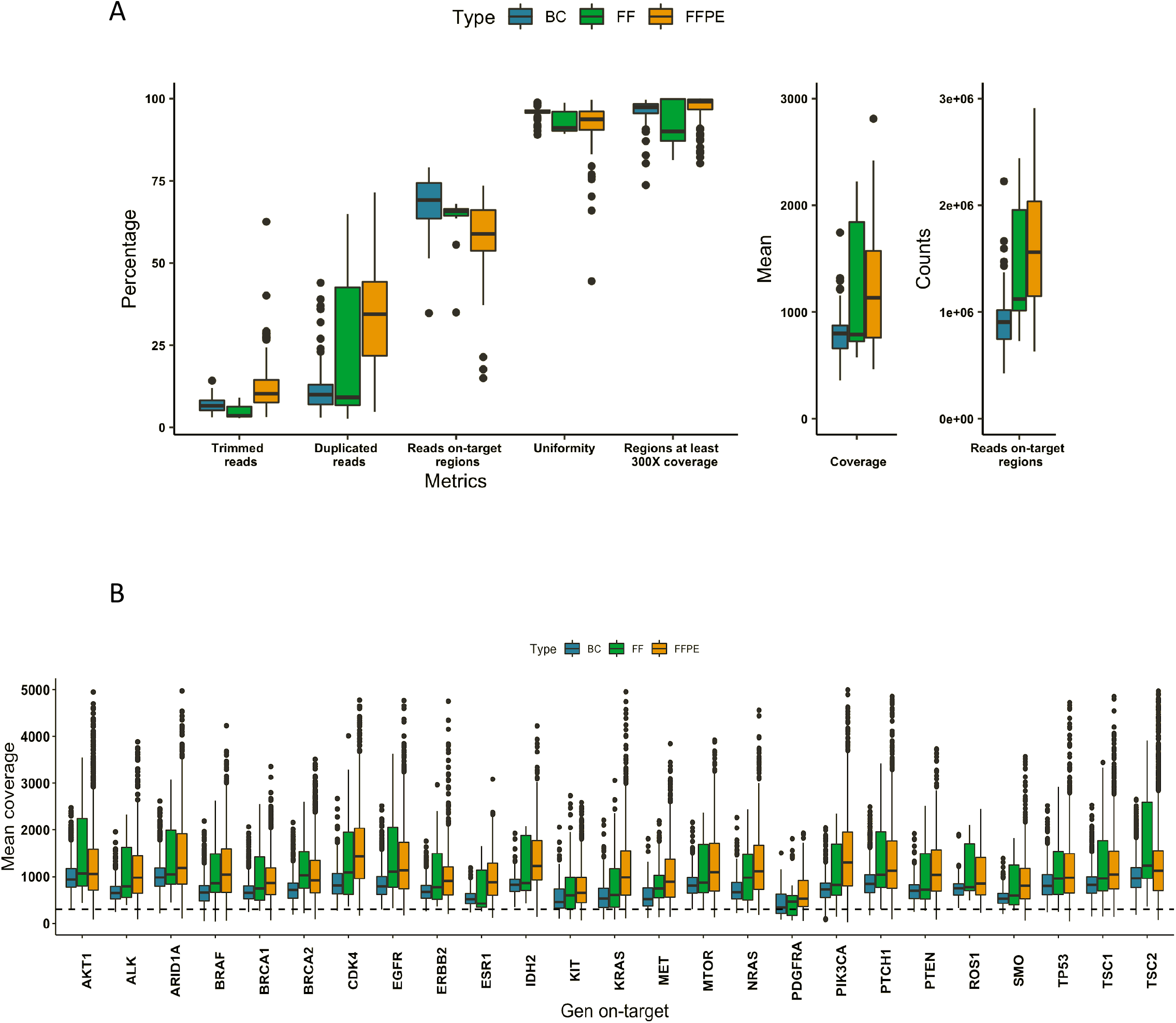
**A)** Box plot of sequencing metrics by type of sample (79 BC, 15 FF, and 108 FFPE). BC: Buffy coat; FF: Fresh Frozen; FFPE: Formalix-fix Paraffin-Embedded. Each box shows the data distribution divided into quartiles. The centerline shows the median, and outlier points are in black. **B)** Box plot of mean coverage of on-target regions per gene for each type of sample: BC (blue), FF (green), and FFPE (orange). The centerline shows the median, and outlier points are shown in black. The vertical dotted line shows 300X coverage.

### Automating bioinformatic analysis and reporting of results

We developed a semi-automated bioinformatic pipeline that starts with raw data from an Illumina sequencer in fastq format, and ends with two reports as observed in Figure 1. First, a technical report with detailed information on data quality, variant calling and annotation is generated. Additionally, a second report containing only information regarding actionable mutations is also created. Human interaction is possible at various stopping points throughout the pipeline to facilitate decisions regarding samples that may need to be repeated or skipped for further analyses. Nevertheless, the entire pipeline can run without human intervention until the technical report. The pipeline is available in a docker image (https://hub.docker.com/repository/docker/labgenomicatumorsec/tumorsec).

### Panel Performance

The panel’s performance was calculated using the reference HD200 (Horizon Discovery) standard FFPE sample containing characterized mutations in the following genes: *BRAF, KIT, EGFR, KRAS, NRAS, PIK3CA, ARID1A, and BRCA2*. As observed in Figure 3A, the assay captured all 13 positive variants. We extrapolated a 0.98 coefficient of correlation (r-squared) with a p-value of 3.221e-10 between expected variant allele frequencies (VAF) from the positive control and those reported by our assay. VAFs ranged from 24.5% to as low as 1%, showing the assay’s high analytical sensitivity.

**Figure 3.**
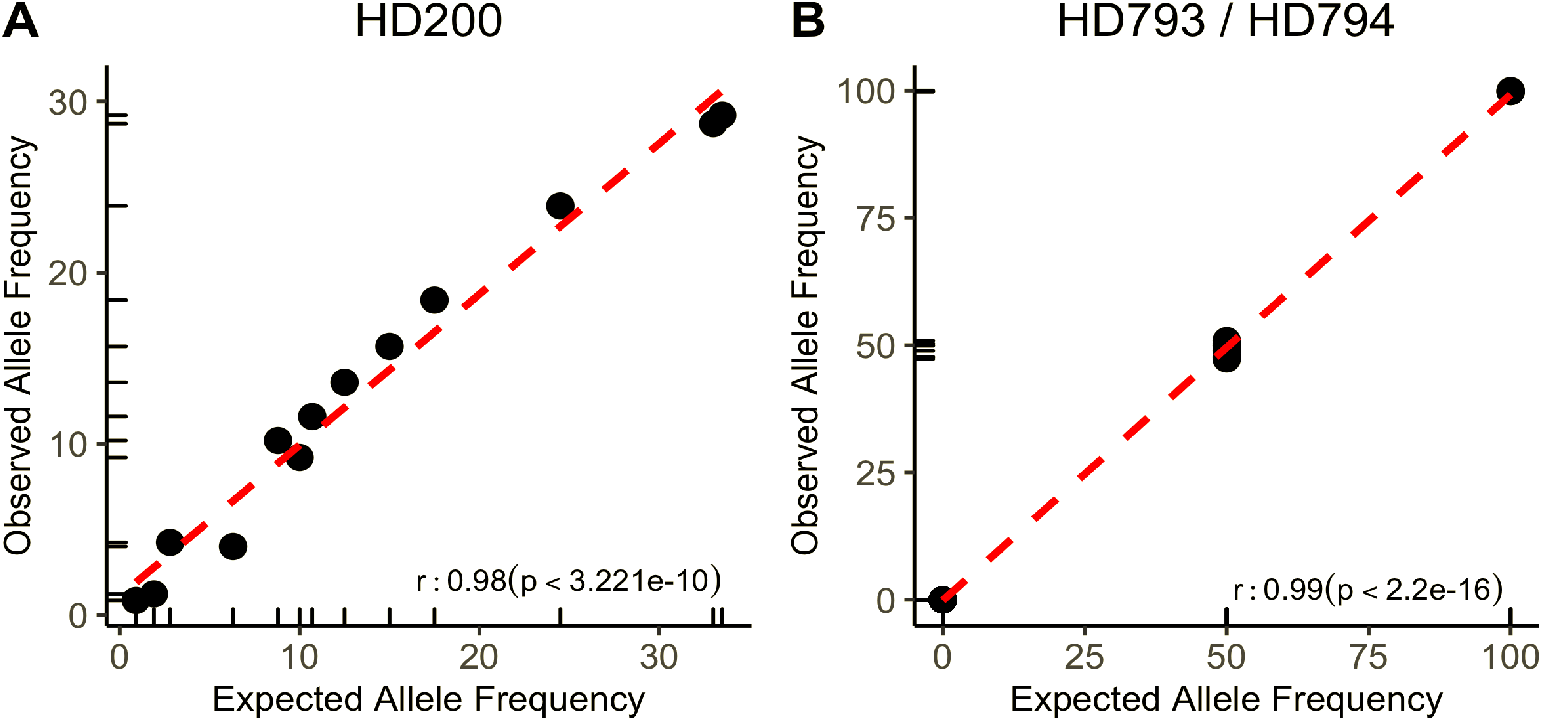
Correlation between expected allelic frequencies for reported variants in the Horizon Discovery controls and those observed by TumorSec. A) FFPE HD 200. B) HD 793 and HD 794.

Additionally, we decided to test the panel’s performance for detecting *BRCA1/2* germline mutations, which are predictive biomarkers for PARP inhibitors therapy in breast, ovarian and prostate cancer (Table 1). Thus, references DNA HD793 and HD794 (Horizon Discovery), which contain known germline variants in *BRCA1* and *BRCA2* at VAF of 50 and 100% were sequenced. Figure 3B shows that 11 out of 11 reported variants were detected at the expected VAF. The correlation coefficient between expected and reported VAF is 0.99 with a p-value of 2.2e-16. Importantly, no mutations were detected in the 15 positions reported as “no-mutated” (true negatives), showing the assay’s high specificity.

For reproducibility assessment, three FFPE samples from different tumors (colorectal, ovary, and gallbladder) were used to prepare two separate libraries each. All samples passed the sequencing metrics threshold with ≥300x coverage in 99.6% of target regions and 89% uniformity. We observed a 100% concordance among non-synonymous variants detected in the different libraries (Figure 4A).

**Figure 4.**
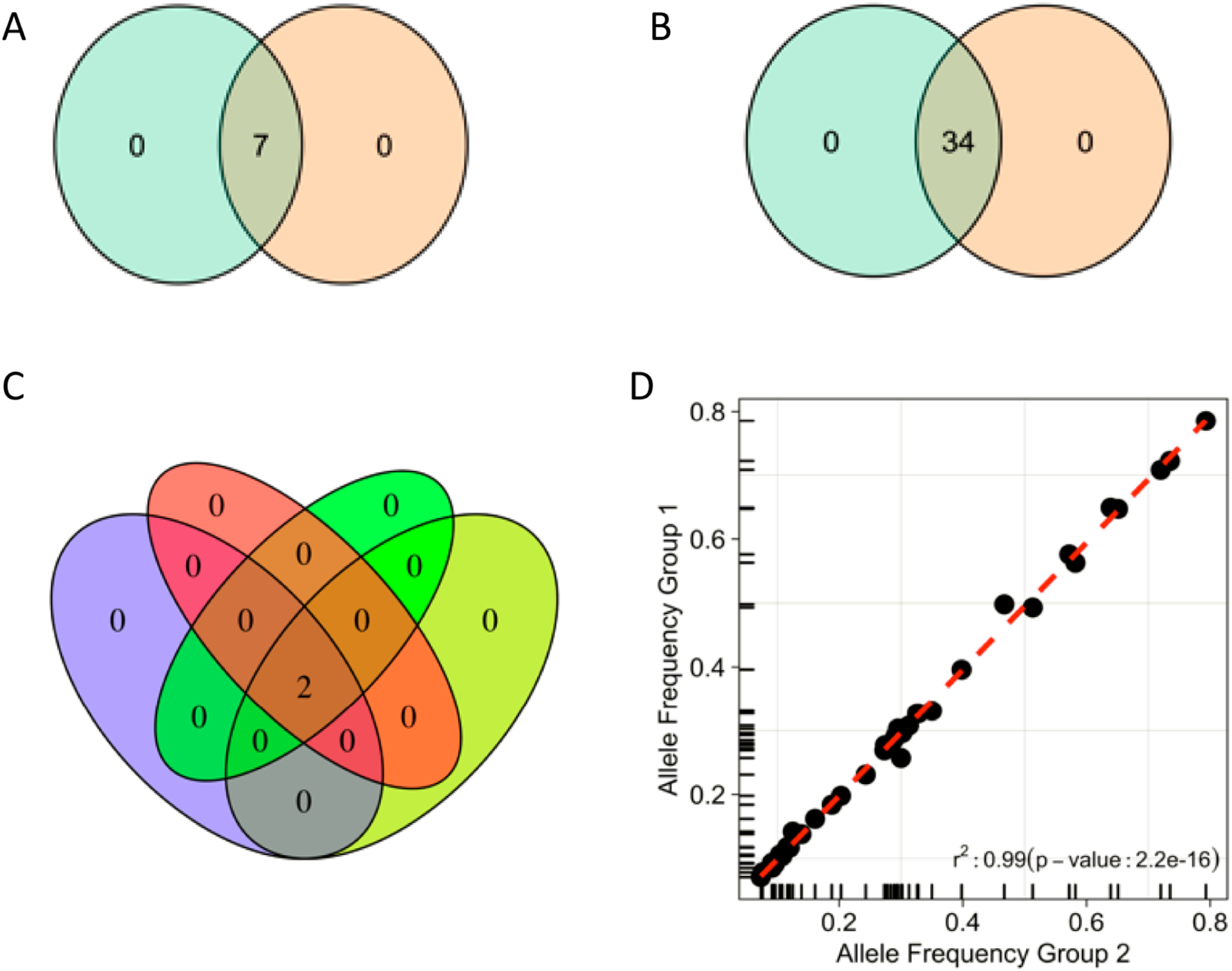
Reproducibility and repeatability. **A)** Within sequencing run reproducibility based on three patient samples, each from a different tumor type (colon, ovaries, gallbladder). Venn diagram displays variants observed in two different library preparations for each of the three samples. **B)** Repeatability between sequencing runs was assessed using the same libraries from three different FFPE cancer and control (HD200) samples in two separate sequencing runs. Venn diagram displays variants observed between two sequencing runs. **C)** One ovarian cancer sample was processed using four different combinations (colors) of library preparation and sequencing runs. **D)** Correlation between allele frequencies of variants obtained in repeatability and reproducibility tests (displayed in A and B).

Inter-runs repeatability was assessed using four FFPE samples (3 ovaries and 1 control). Different libraries were sequenced in different runs. Reproducibility of sequencing metrics (94% of target regions with ≥300x coverage and ≥87% uniformity) and concordance of detected variants were also observed (Figure 4B). One ovarian FFPE sample was assessed in three different library preparations and three separate sequencing runs (Figure 4C). A high correlation (r=0.99) was observed between VAFs called in all the different settings (Figure 4D).

### Comparison between FFPE, Fresh frozen and blood gDNA

In order to assess whether the protocol and bioinformatic workflow for detecting somatic mutations discard FFPE-induced artifacts and germline variants, we sequenced FFPE, fresh frozen tumor (FF) and blood (buffy coat) samples (triad) from six ductal breast carcinoma subjects. For tumor samples, biopsies used presented <5% necrosis and a 42-80% cellularity.

Variants detected among all sample triads for each of the six subjects are outlined in Figure 5. We reported 9 variants in the FF sample set, 7 of these variants were also reported in the matching FFPE samples. It is worth noting that the 2 variants detected in a FF sample (FA6-005) were found in the FFPE sample but at frequencies <5% (the LOD established for the assay). To further explore FF and FFPE samples’ concordance, we plotted the allele frequencies of both synonymous and non-synonymous variants detected in both sample types (Figure 6). Variants with AF<5%, display a low r-value (0.68, p-value of 2.479e-09). However, when all variants (73) are analyzed, correlation increased (r=0.95, p-value of < 2.2e-16). Importantly, no germline variants and no variants exclusive for FFPE samples were detected using the pipeline for somatic mutations.

**Figure 5.**
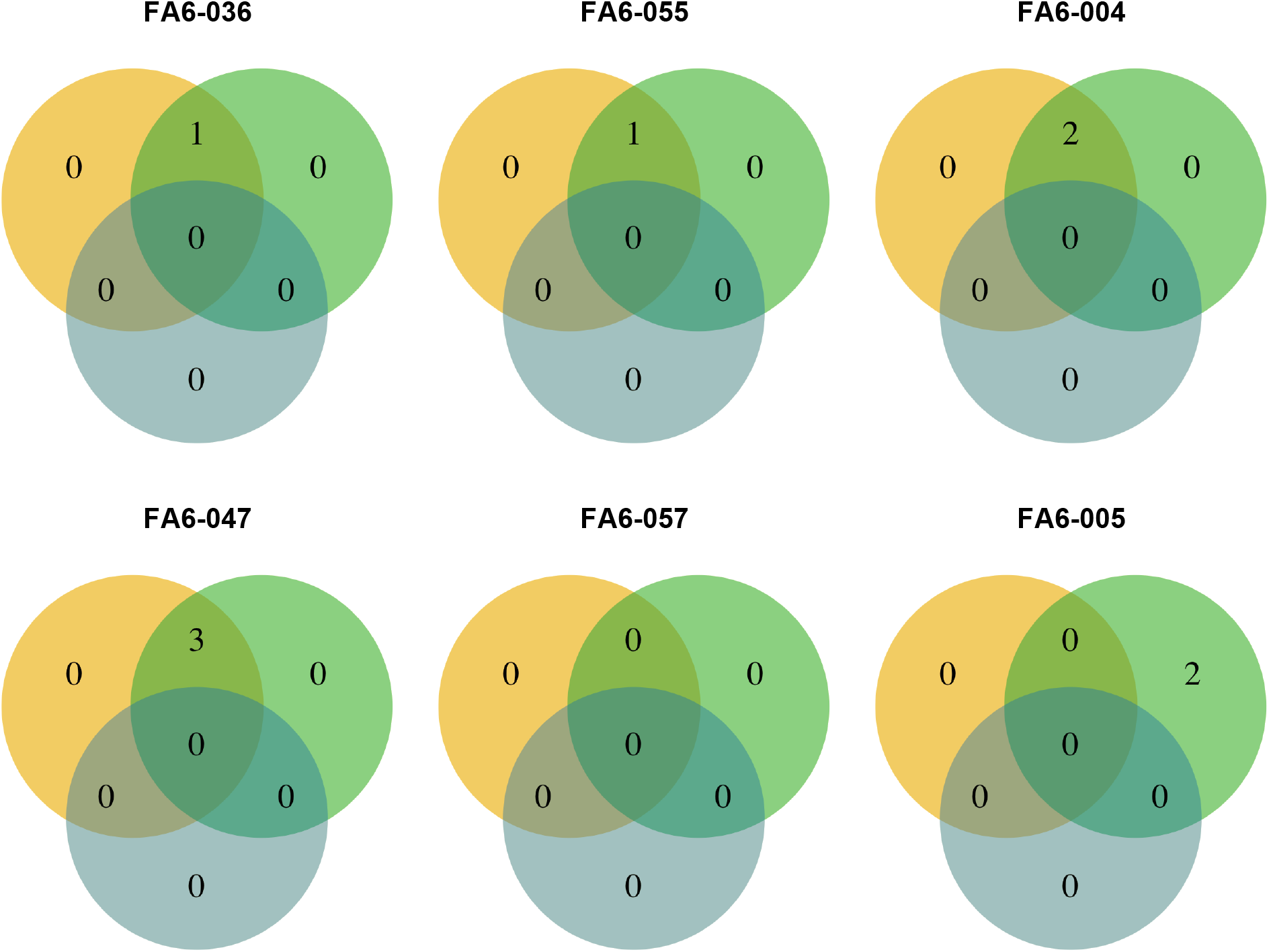
Venn diagram outlining non-synonymous variants reported for six different patients diagnosed with ductal breast carcinoma among three different sample types for each: FFPE (orange), Fresh Frozen (green), blood (blue).

**Figure 6.**
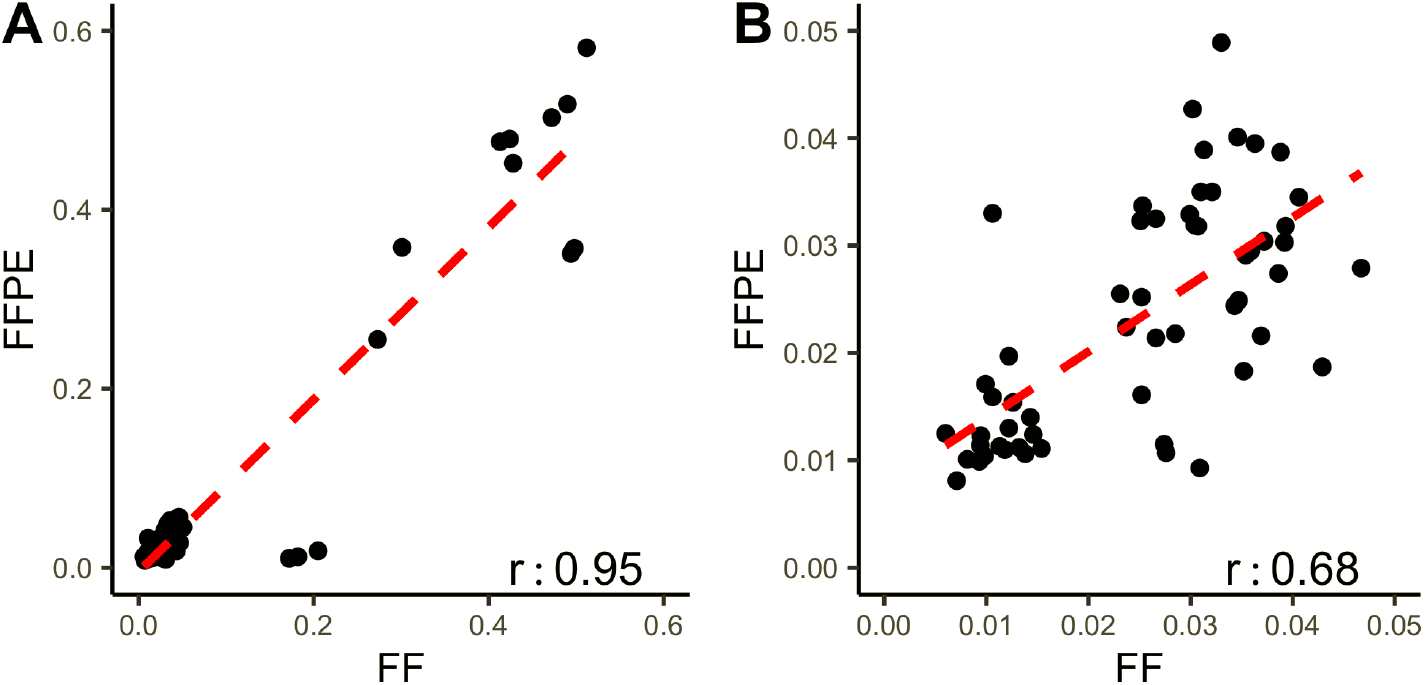
Synonymous and non-synonymous variant’s frequency comparison between FFPE and Fresh Frozen samples. A) Correlation between allele frequencies of all variants (0-60%) found in FFPE and Fresh Frozen samples. B) Correlation between frequencies of variants with VAF (below LOD).

### Validation of the assay in clinical samples

To validate the assay and analysis capabilities in “real world” samples, we processed 176 tumor biopsies from different clinical sites. 123 out of the 176 were successfully sequenced (108 FFPE and 15 FF): breast (14), ovary (69), gastric (1), gallbladder (23), and colorectal (16).

All variants with allelic frequency over 1% reported in at least one of the following four germline population variant’s databases (PVDs): GnomAD, ESP6500, ExAC, and 1000 Genomes, were eliminated. However, given that the Latin American population is not well represented in these repositories, somatic mutations were initially overestimated due to their absence or low (<1%) AF. Thus, for the classification of remaining variants, we used the algorithm depicted in Supplementary Figure 2, based on annotations in COSMIC, dbSNP, CLINVAR, and PVDs databases; and recommendations from Sukhai et al. (2019)^23^. Filtering was extended to include all under-represented populations that have information in GnomAD and ExAC. Additionally, a local repository was built using germline data from current and other related works. The resulting variants were classified as Germline, Somatic, Putative Germline, Putative Somatic, Putative Germline Novel, or Putative Somatic Novel.

We found a total of 245 protein affecting variants in the 123 samples. Among these, 187 non-synonymous somatic and putative known and novel somatic variants were identified in 105 out of the 123 samples (85%) (Table 3). A hundred thirty-eight (138) were unique variants. Figure 7 shows a breakdown of these mutations by variant and tumor type. Among the breakdown of these variants are deletions and insertions (including those causing frameshifts), missense and nonsense mutations, and variants positioned in splice sites. As depicted in Figure 7A, missense mutations are the most prevalent types of mutation among all samples. Samples from ovarian tumors are the most abundant in this study and have the highest number of unique variants (Figure 7B).

**Table 3.**
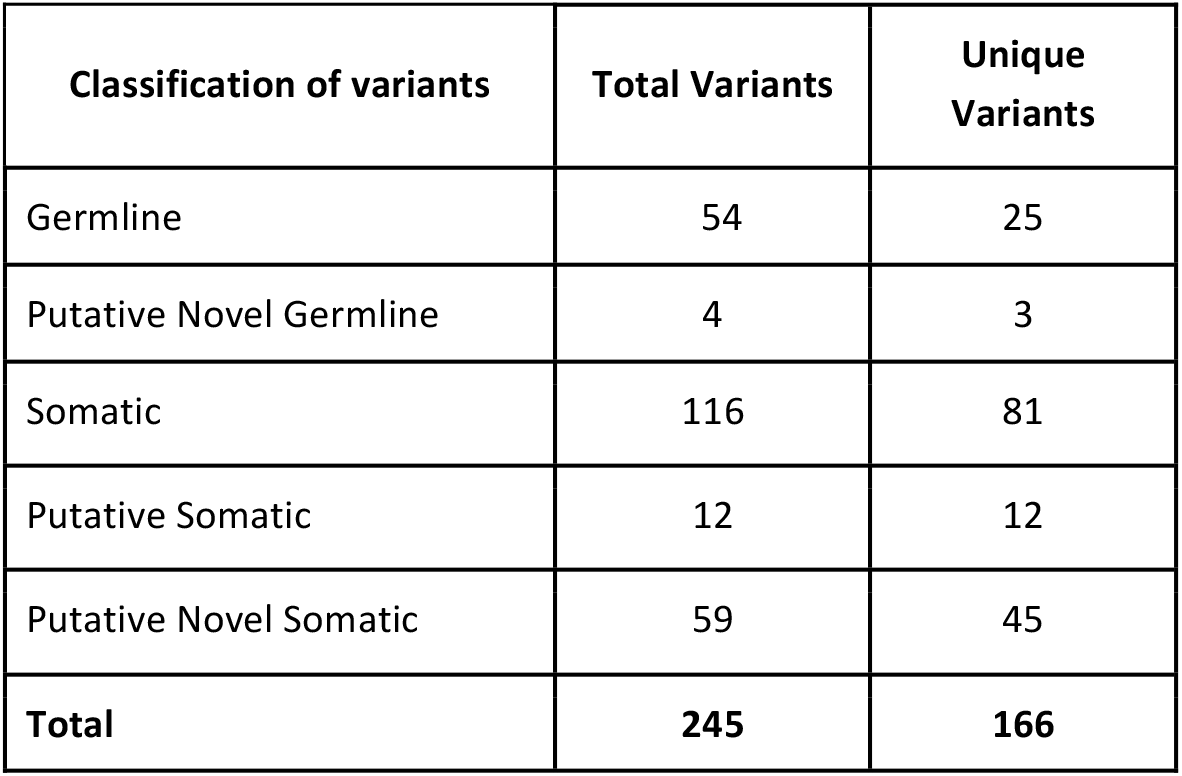
Classification of non-synonymous variants in 123 quality passed tumor samples. It is shown the number of total and unique variants by classification.

| Classification of variants | Total Variants | Unique Variants |
| --- | --- | --- |
| Germline | 54 | 25 |
| Putative Novel Germline | 4 | 3 |
| Somatic | 116 | 81 |
| Putative Somatic | 12 | 12 |
| Putative Novel Somatic | 59 | 45 |
| **Total** | **245** | **166** |

**Figure 7.**
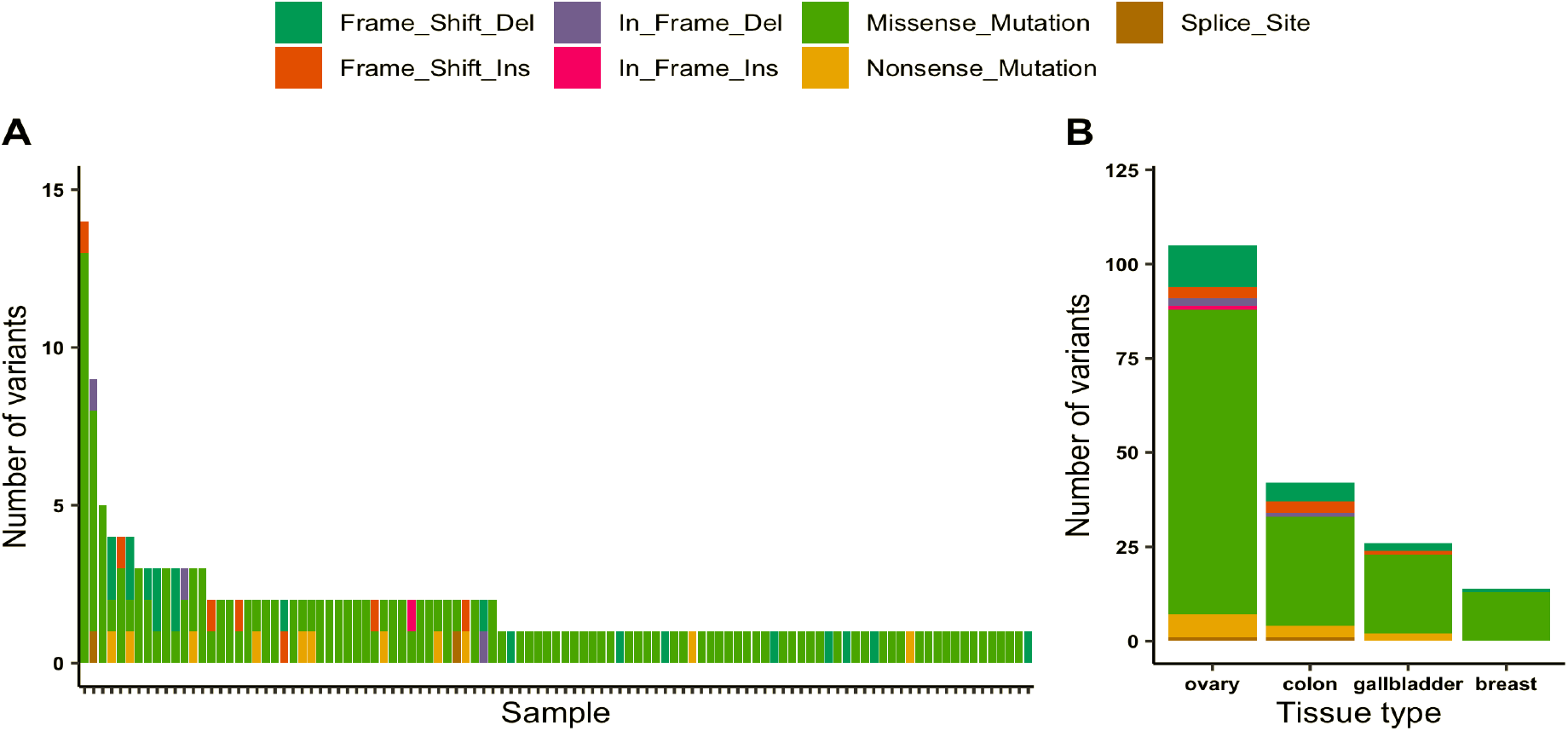
Non-synonymous Somatic Variants detected across all tumor samples. 187 non-synonymous somatic variants were detected in 105 out of the 123 samples, including “putative known somatic” and “putative novel somatic” variants. A) Number of detected variants per sample and mutation type. B) Number of unique variants somatic variants per tumor type.

### Identification of Biomarkers for Targeted Therapies

131 (70%) out of the 187 identified somatic variants are described as a biomarker for drug response, supported by different levels of evidence: FDA guidelines (43), NCCN guidelines (9), late trials (37), early trials (116), case report (42), and (108) pre-clinical data. The affected gene and target drug associations with supporting evidence from “case reports” to “FDA guidelines” are depicted in Figure 8, where is outlining: (1) The fraction of samples with reported genetic alterations, (2) Level of existing evidence for the biomarker, (3) Gene affected and (4) the drug association (resistant or responsive). Table 4 contains a detailed description of the biomarker mutations supported by FDA and NCCN clinical guidelines found in this study.

**Figure 8.**
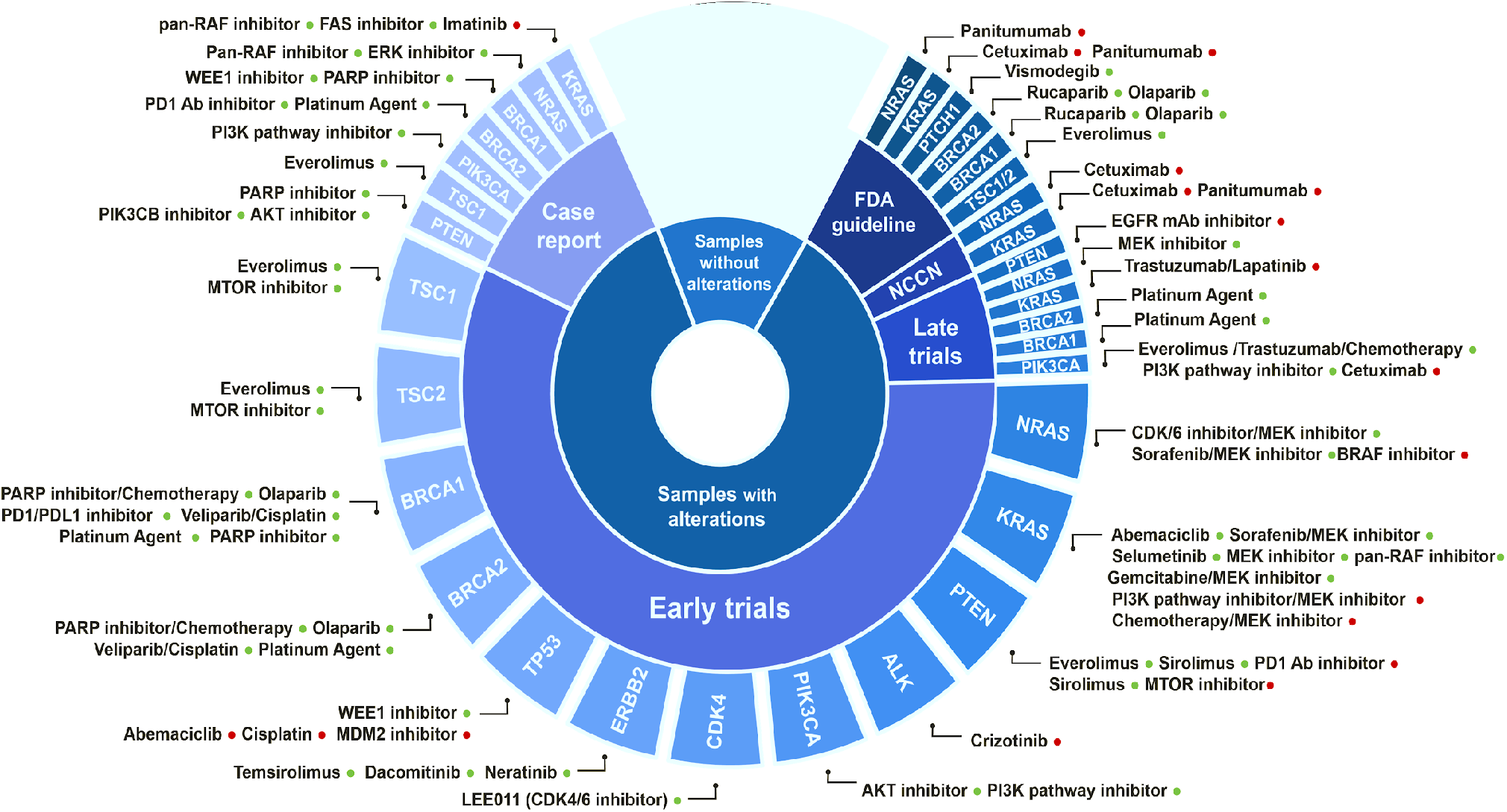
Predictive biomarkers for therapy response identified in the analyzed clinical tumor samples. The biomarker gene, the associated drugs, and the level of evidence supporting this association in solid tumors are depicted. Positive (drug-responsive) and negative (drug resistance) associations are indicated by green and red dots, respectively.

**Table 4.**
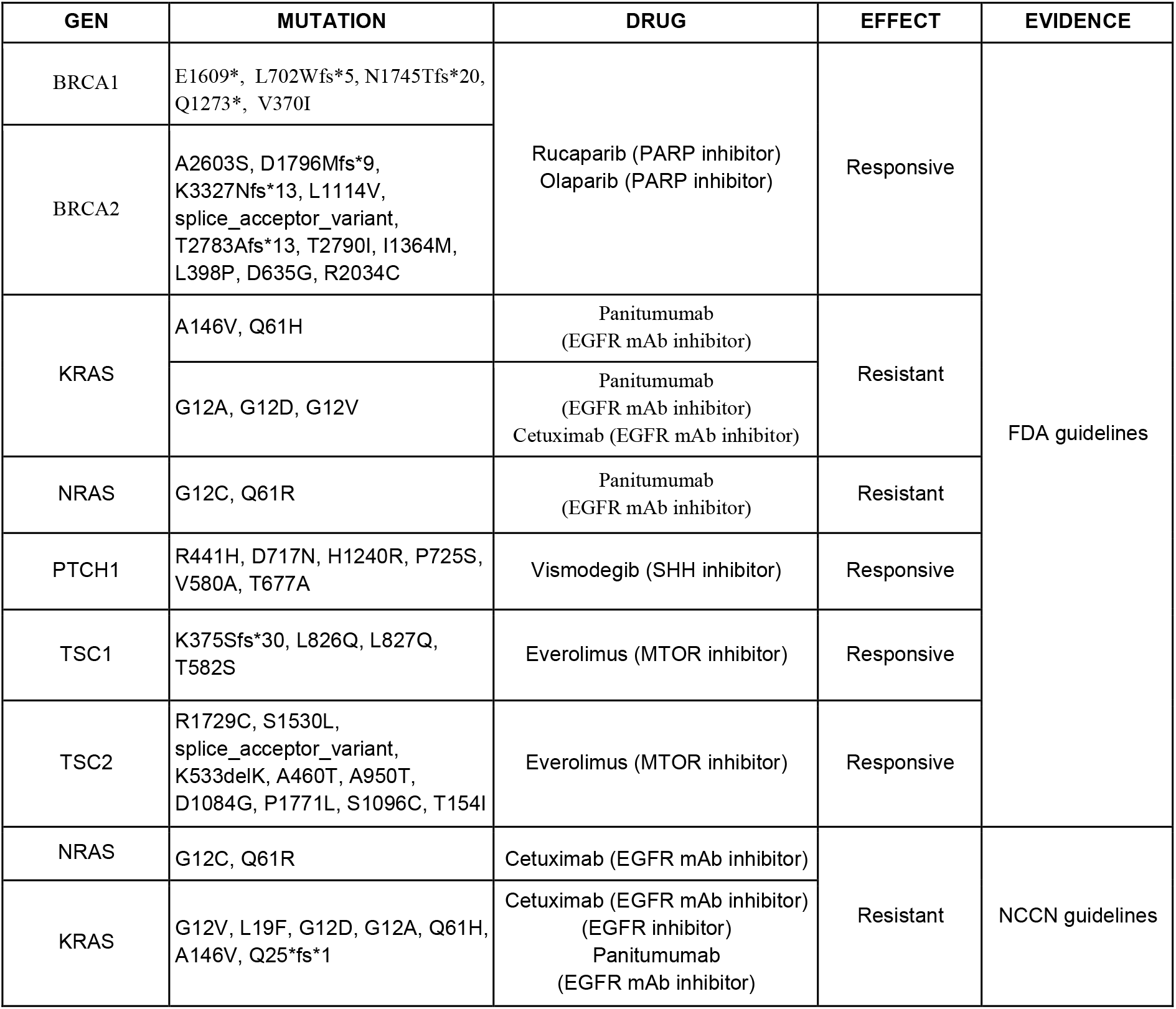
Biomarker mutations supported by FDA (43) and NCCN and other clinical guidelines (9), found in the analyzed clinical samples. The Associated drug, the mutation’s predictive effect and supporting level of evidence are indicated.

## Discussion

As a result of its global adoption and implementation, the clinical utility of NGS in the field of oncology has a rapidly growing body of evidence. The ability to get massive amounts of genetic information from small amounts of tissue provides clear advantages for decision-making against cancer^37,4,2^. Nevertheless, a large portion of cancer patients around the world do not have this option readily available. This work attempts to favor the implementation of NGS in the Latin American health system by showcasing a locally-developed assay, accompanied by an open-source automated analysis focused on the target population’s needs.

A critical consideration for implementing NGS in low resource settings is finding a workflow compatible with low-quality, highly degraded samples. Although fresh frozen tissue is the gold standard for molecular analyses, its use in clinical practice is impractical because of its high cost and technical difficulty. The sample storage infrastructure found in the developed world, with dedicated −80C and −20C freezers, is often not in the budget for many Latin American diagnostic laboratories. FFPE tissue samples are much more cost-effective as they can be stored at room temperature. However, tumor biopsies in this region are often fixed with formalin with different protocols and laboratory environments, producing varied DNA damage during and after the formalin fixation process (e.g., fragmentation, degradation, crosslinking^24,38^).

DNA quality is affected by the type of formalin used for tissue fixation and the time since preservation^25^, both of which vary highly in laboratories across Chile and Latin America. A total of 49 FFPE samples failed to pass the DNA, library, or sequencing quality controls. Most of the failed samples are colorectal (33/40) and gallbladder (13/36) samples. Twenty-four failed colorectal samples had low on-target rates, suggesting issues with hybridization and or capture, while the 13 gallbladder samples did not pass the library preparation quality metrics.

In general, FFPE samples showed a higher percentage of duplicates and off-target reads (Table 2). However, these characteristics do not affect sequencing results. FFPE samples have the highest mean on-target region’s coverage compared to FF and BC. Removing duplicates is intended to reduce noise during the variant identification process and minimize false positives. Our results suggest removing duplicates has little effect on this panel’s performance at the sequencing depths we are interested in (∼300X). As sequencing technologies continue to advance, PCR duplicate removal will become less of an issue^26^.

Somatic mutation analysis is critical in cancer research. Currently, there is no community consensus about the most appropriate variant caller for somatic mutations^27^. For this reason, we incorporated six variant callers capable of producing highly accurate somatic mutation calls for both SNVs and small Indels. Nevertheless, when comparing the frequency of mutations per gene found in our cohort with the reported in international databases, a higher frequency in some of the genes was evident. Somatic variant callers discard germline variants by interrogating the reference genomes and population databases such as 1,000 Genomes, where Latin American genetic variation is not well represented. Thus, we implemented a more accurate bioinformatic pipeline that allows variants’ classification as somatic, germline, and putative somatic/germline. This variant calling process highlights the extra layer of difficulty Latin American researchers and clinical laboratories face due to the absence of reference genomes representative of our population in the major databases. Overestimation of somatic variants is a problem when facing the tumor of a patient from any region or ancestry without a reference genome informative of the genetic variation in that specific population. This issue is critical for therapy determinants such as the tumor mutational load, which should be carefully interpreted in these patients^39^.

The best approach for resolving the somatic vs. germline mutations issue is to include respective blood samples alongside biopsies. However, this raises the assay’s cost per patient, which may delay the assay’s implementation.

To achieve more accurate somatic variant calling, we need further efforts towards genetic characterization of the Latin American and other under-studied populations. Building an inclusive tumor reference genome database will allow discovering novel somatic mutations and non-explored correlations to the disease.

## Data Availability

The bioinformatic pipeline, user manual, and tutorials will be available after publication in the GitHub repository called Pipeline-TumorSec (https://github.com/u-genoma/Pipeline-TumorSec).
The pipeline is available in a docker image (https://hub.docker.com/repository/docker/labgenomicatumorsec/tumorsec).

https://hub.docker.com/repository/docker/labgenomicatumorsec/tumorsec

## Acknowledgments

This work was funded by grant FONDEF IDEA IT16I10051. The HPC computational resources used in this work were funded by grants VID INFRAESTRUCTURA 0440/2018 (University of Chile), FONDEF D10E1007 and D11I1029 (CONICYT), and VID U-Redes 704/2016 (University of Chile), FONDECYT Fondo Nacional de Investigación y Tecnología 1171463. Probes and other consumables were partially provided by Roche Sequencing Solutions (Pleasanton, CA). We thank Daniela Diez and Vania Montecinos for clinical coordination, and the personnel from “Biobanco de Tejidos y Fluidos de la Universidad de Chile” for their permanent support. We also thank all patients who altruistically donated their samples to this research.

