## Supplementary Figures for "Validation of a NGS panel, with automated analysis, designed for detection of medically actionable tumor biomarkers for Latin America"

A

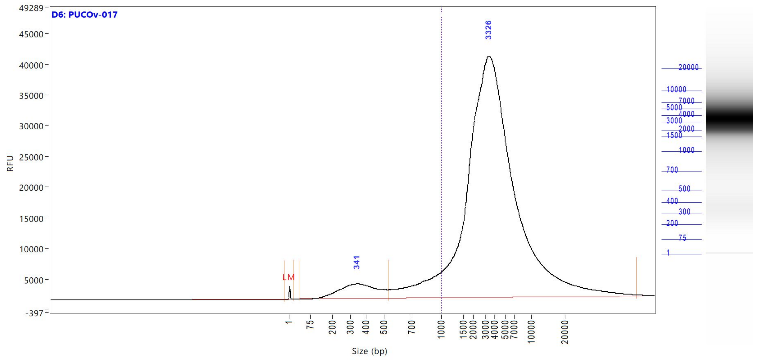


B

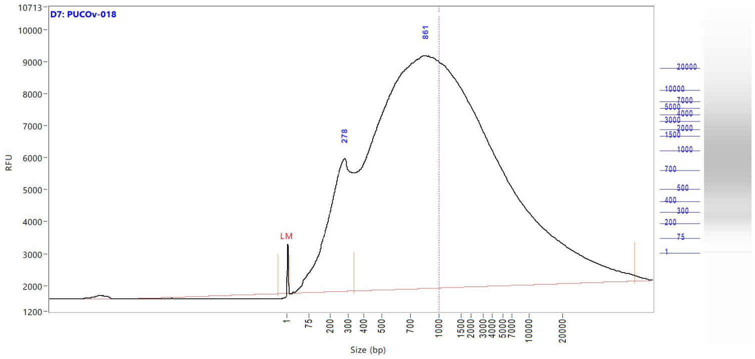


C

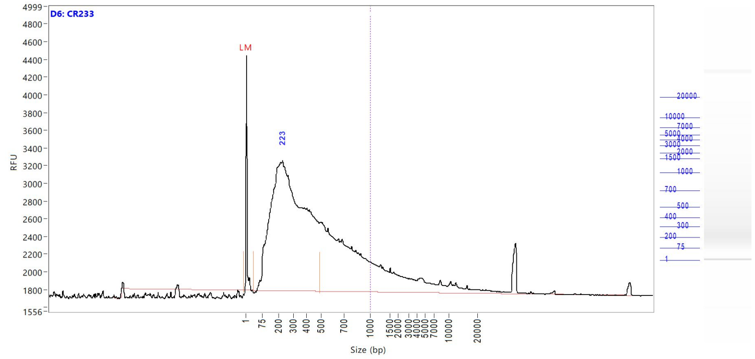


D

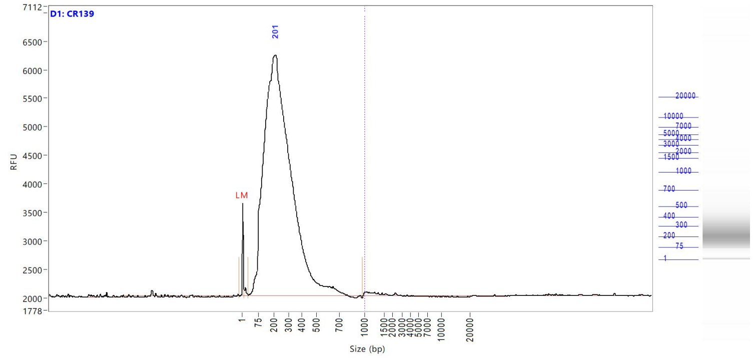


**Supplementary Figure 1.** Wide range of integrity of DNA extracted from FFPE tissues. Examples of FFPE DNA's electropherograms obtained by the Fragment Analyzer Automated CE System indicating the range of degradation with a size threshold of 1000bp (dotted line) using HS Genomic DNA Analysis Kit (DNF-488). **A)** Undegraded FFPE DNA (>1000bp). **B)** Moderately degraded FFPE DNA (>400-1000bp). **C)** Degraded FFPE DNA (<400bp). Due to their excessively degraded nature, low-quality samples with less than 200bp **(D)** are not recommended for use with the Tumorsec workflow.

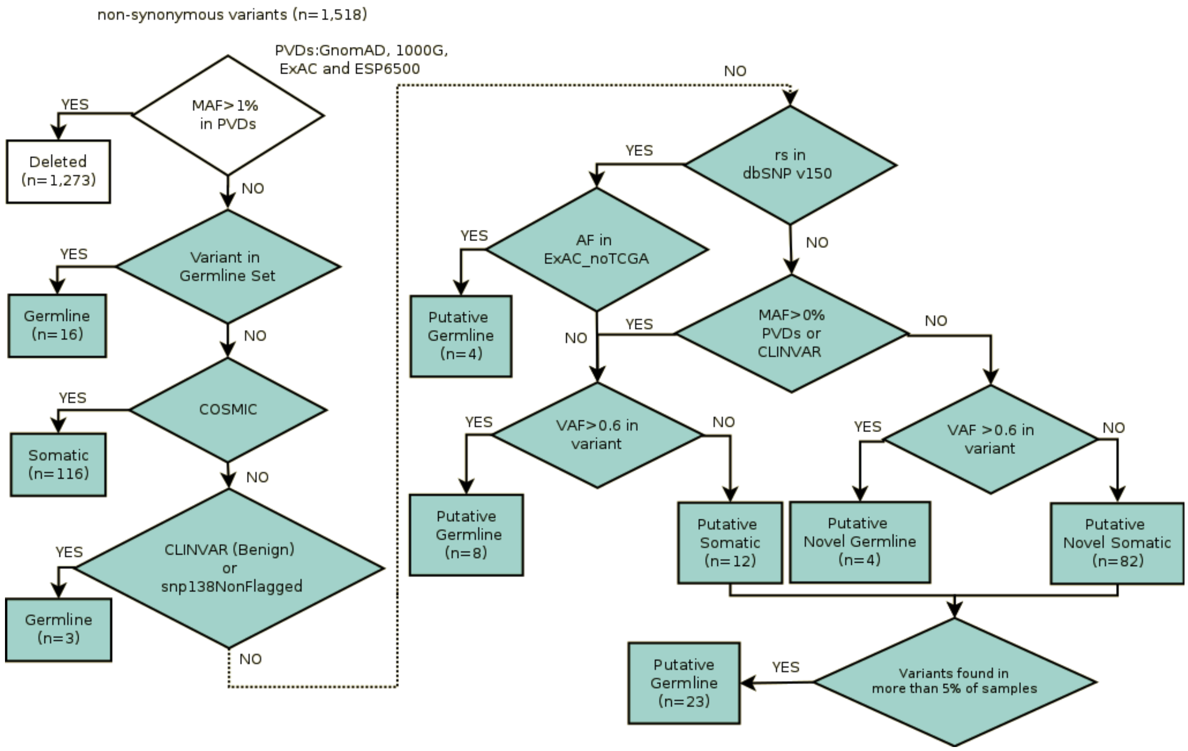


**Supplementary Figure 2:** Variant classification algorithm. This image shows our flow diagram, which utilizes nine conditionals. Each conditional sequentially classifies variants as somatic or germinal (known, putative, or putative novel). 1) Variants with MAF over 1% in PVDs are deleted (n=1,273). 2) variants found in blood samples (BC) are labeled as germline in tumor samples (n=16). 3) Variants with a COSMIC annotation are labeled as somatic (n=116). 4) Benign or likely benign variants according to Clinvar or rs in snp138NonFlaged, are labeled as putative germline (n=3). 5) Variants with rs in dbSNP and reported in ExAC no-TCGA are labeled as putative germline (n=4). 6-7) Variants without classification and rs in dbSNP, MAF between 0-1% in PVDs or Clinvar information, are labeled as putative germline (n=8) or putative somatic (n=12) when VAF in the samples is >60% or <60%, respectively. 8) Variants without classification are labeled as putative novel germline (n=4) or putative novel somatic (n=82) when VAF in the samples is >60% or <60%, respectively. 9) Finally, variants classified as putative somatic or putative novel somatic found in more than 3 (5%) samples are re-classified as putative germline (n=23). MAF: minor allele frequency, PVDs: Population Variants Databases, COSMIC: Catalogue of Somatic Mutations in Cancer, Snp138NonFlaged: dbSNP without flagged SNPs (SNPs with MAF < 1% or unknown, mapping only once to reference assembly or with a clinical association, rs: Reference SNP, VAF: Variant Allele Frequency.
